# From Slices to Volumes: A Scalable Pipeline for Developing General-Purpose Brain MRI Foundation Models

**DOI:** 10.1101/2025.04.12.25325728

**Authors:** Feng Su, Xiaoping Yi, Ye Cheng, Yongjie Ma, Wenqiang Zu, Qing Zhao, Gengdi Huang, Lei Ma

## Abstract

Foundation models exhibit a remarkable capacity in extracting subtle features from brain MRI, demonstrating transformative potential for the precise diagnosis of brain diseases. Here, we introduce BrainMRIFM, a scalable pipeline for developing both slice and volume brain MRI foundation models that achieve computational efficiency alongside powerful representational capabilities. BrainMRIFM utilizes a novel slice-to-volume training paradigm: a slice model is initially pretrained for MRI slice representation, then its parameters are transferred to the corresponding volumetric model. The models were trained on an unprecedented dataset comprising 140,501 multi-modal MRI volumes from 42,297 subjects, aggregating data from 130 public datasets and 8 custom clinical centers. Our innovative in-house datasets contribute over 80% of the total patient cases for seven major brain diseases that lack open-source MRI data. Pretrained on 25 million high-quality MRI slices, the three slice foundation models demonstrated consistent performance improvements across all three downstream tasks, achieving a maximum accuracy improvement of 10.49% compared to ImageNet-initialized models. The SimMIM-SwinT volume MRI foundation model, building upon the high-performance slice foundation model, exhibited robust performance across seven brain tumor diagnostic tasks, with a maximum AUROC improvement of 12.19% compared to conventional ResNet50-based task-specific models. Additionally, attention maps and saliency visualizations confirmed the model’s capability to accurately localize pathological features. The BrainMRIFM pipeline and associated resources represent a significant advance toward developing brain MRI foundation models, with clear potential for extension to other neuroimaging applications.

## Introduction

Magnetic resonance imaging (MRI) offers non-invasive checking of three-dimensional (3D) details with superior soft tissue contrast, making it an essential radiological imaging technology for neuroscience research and brain diseases diagnoses^1–3^. Deep learning (DL) models have demonstrated remarkable capabilities in identifying subtle pathological features from complex MRI data, thereby holding great promise for intelligent diagnostics^4,5^. However, most of these task-specific DL models often exhibit limited generalizability, and their reliance on large amounts of annotated data presents significant barriers to widespread application. Foundation models, developed through pretraining and fine-tuning, represent the forefront of advancements in artificial intelligence. These models utilize self-supervised learning (SSL) to automatically extract meaningful representations from vast amounts of unlabeled data and can be fine-tuned to perform a wide range of downstream tasks^6^. Radiological imaging foundation models have already achieved significant success on various imaging modalities (e.g., MRI, CT and 3D OCT) across different organs (e.g., lungs, livers, and heart)^7–9^. However, brain MRI presents unique characteristics, including lower texture and contrast, complex imaging sequences, and relatively limited data availability. As a result, developing a general-purpose brain MRI foundation model remains a significant challenge.

Contrastive learning and image reconstruction are two fundamental paradigms in self-supervised learning, enabling foundational models to achieve meaningful MRI representation learning. BrainIAC and AnaCL are typical brain MRI foundation models based on contrastive learning. BrainIAC, a general-purpose model built on 3D ResNet50, was developed using 48,519 multimodal brain MRI volumes sourced from 35 datasets across 10 neurological conditions, and showed great performance in MRI sequence classification, brain age prediction, mutation classification, and overall survival prediction^10^. Similarly, AnaCL, an anatomy-oriented model based on 3D ResNet18, was developed using 21,155 T1w brain MRI volumes from 7,908 individuals, exhibiting significant potential in various diagnostic classification and clinical assessment score prediction tasks^11^. BrainSegFounder and MoME are exemplary segmentation foundation models based on image reconstruction techniques, including Encoder-Decoder and masked image modeling (MIM). BrainSegFounder, built upon the 3D SwinUNETR^12^, was pretrained using 82,800 paired T1w and T2w MRI volumes, followed by self-supervised re-pretraining and supervised fine-tuning on downstream datasets, demonstrating great performance in brain tumor and stroke lesion segmentation^13^. MoME segmentation model utilized the nnU-Net architecture and was developed using 6,585 annotated 3D MRI volumes. MoME employed multiple expert networks to process different MRI modalities, showcasing its potential for automated segmentation of various brain lesion types across diverse MRI modalities^14^. Furthermore, Vision Transformer-based brain tumor (ViTBT) foundation models, trained on 57,621 MRI volumes using MIM techniques, have exhibited improved performance in tumor discrimination and molecular status prediction compared to traditional convolutional neural network models^15^. The integration of medical prior knowledge^16–18^ and contrastive loss^19^ during the pretraining process has been empirically validated to enhance the efficacy of representation learning. Notably, the BME-X foundation model innovatively combines an automated tissue classification network with an image enhancement network to improve image quality. Developed using 2,448 synthesized corrupted images and 10,963 T1w MRI slices from 19 public datasets, this model has shown substantial potential in enhancing downstream tasks, including segmentation, registration, and diagnostic applications^20^. Additionally, segmentation foundation models trained on diverse medical image datasets have demonstrated strong performance in brain tumor segmentation tasks^21,22^.

These advancements have confirmed the excellent performance and strong generalizability of brain MRI foundation models. However, compared to pathological foundation models and natural image or video foundation models, most of these studies suffer from a significant lack of scale and diversity in brain MRI datasets, which hinders the comprehensive training of foundational models. Furthermore, the direct pretraining of 3D models not only poses challenges in representation learning and computational efficiency but also exacerbates the issue of the limited availability of volumetric brain MRI data.

In this study, we propose BrainMRIFM, a scalable pipeline for developing general-purpose brain MRI foundation models (Fig. 1). Our work presents three main contributions. First, the largest and most diverse brain MRI dataset to date. We curated an unprecedented collection of 140,501 multi-modal MRI volumes from 42,297 subjects, aggregating data from 130 public datasets and 8 custom clinical centers. This dataset includes healthy subjects as well as individuals with over 22 distinct major brain diseases. Our novel in-house datasets contribute more than 80% of the total patient cases for seven major brain diseases that lack open-source MRI data. This substantial addition dramatically enhances the disease coverage and diversity of existing brain MRI resources. Furthermore, we established an MRI slice dataset comprising 25 million high-quality slices, following rigorous data cleaning and quality control procedures. Second, a scalable pipeline for developing slice-to-volume MRI foundation models. BrainMRIFM initially pretrains slice foundation models using self-supervised learning (SSL) methods, enabling the models to learn effective MRI representations and extract critical features from the MRI slices. Subsequently, the parameters of the pretrained slice models are transferred to the corresponding volume model, enabling powerful volume feature extraction and enhanced practical applicability. This scalable slice-to-volume paradigm simultaneously achieves computational efficiency and powerful capability. Third, the slice and volume foundation models exhibited significantly enhanced diagnostic performance. Three slice foundation models (SimMIM-SwinT, MoCo v3-ViT, MAE-ViT) demonstrated consistent performance improvements across all three downstream tasks, achieving a maximum accuracy improvement of 10.49% compared to ImageNet-initialized models. The SimMIM-SwinT volume MRI foundation model, building upon the high-performance slice foundation model, exhibited robust performance across seven brain tumor diagnostic tasks, with a maximum AUROC improvement of 12.19% compared to conventional ResNet50-based task-specific models. Furthermore, attention maps confirmed the model’s ability to accurately localize pathological features.

**Fig. 1.**
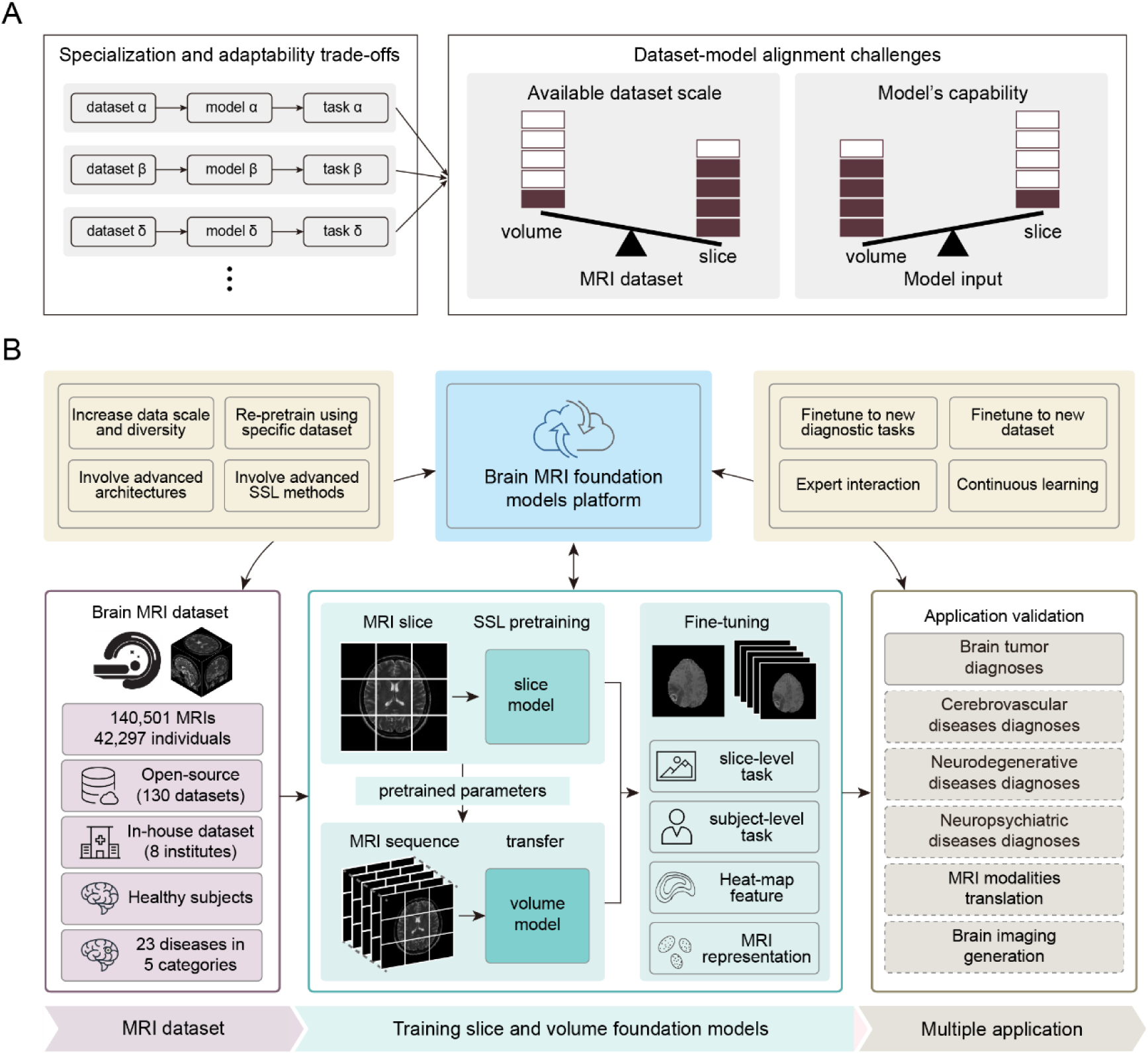
Objectives and flowchart of the BrainMRIFM framework. (A) Objectives of BrainMRIFM. Left, to address the limitations of specialized small models in generalizability, BrainMRIFM aims to develop brain MRI foundation models that achieve optimal trade-offs between specialization and adaptability. Right, BrainMRIFM specifically targets dimensional constraints in modeling prrocess, overcoming significant challenges related to dataset-model alignment. (B) Flowchart of BrainMRIFM. The pipeline begins with the curation of a large-scale, diverse brain MRI dataset. Subsequently, it establishes both slice and volume MRI foundation models for brain disease diagnosis. Designed as a modular yet integrated system, BrainMRIFM enables targeted optimization of individual components while maintaining overall architectural coherence.

## Results

### Pipeline for brain MRI foundation models

The foundation model holds immense potential to unlock the capabilities of MRI for the diagnosis of brain diseases. As a generalist approach, the foundation model can overcome the limitations of task-specific models, which often struggle to meet the diverse demands of clinical applications (Fig. 1A). However, during the development of foundation models, the misalignment between the characteristics of slice and volume datasets and the model architecture remains a significant challenge (Fig. 1A). Our primary objective is to develop a highly efficient and generalizable MRI foundation model that integrates the respective advantages of slice and volume modeling. This model will be capable of diagnosing a broad spectrum of diseases and adaptable to data from various medical centers (Fig. 1B).

To the best of our knowledge, our study represents the largest and most diverse brain MRI dataset reported to date, comprising 140,501 multi-modal MRI volumes from 42,297 individuals (Fig. 1B). This dataset was meticulously curated by integrating nearly exhaustive influential open-source brain MRI from 130 public datasets (Supplementary Material Ⅰ). Additionally, we included a custom dataset collected from 8 hospitals in China. The dataset demonstrates remarkable diversity in terms of diseases, encompassing healthy subjects as well as individuals with over 22 distinct major brain diseases. Following rigorous data cleaning and quality control procedures, we extracted 25 million high-quality MRI slices to pretrain a foundation model, all of which were validated using the perception-based image quality evaluator (PIQE) to ensure superior image quality. Subsequently, we established both pretrained slice and volume MRI models (Fig. 1B). The slice model was pretrained on 25 million unlabeled MRI slices using self-supervised learning (SSL) methods, enabling the model to learn effective MRI representations and extract critical features from the MRI slices. The parameters of the pretrained slice model were then transferred to the volume model, enhancing its capability to extract efficient volumetric MRI features. During the application phase, the pretrained foundation models can be fine-tuned with few labeled data, facilitating their deployment for a variety of diagnostic tasks at both the slice-level and subject-level. Here, we have successfully applied the established brain MRI foundation model to various typical diagnostic tasks for brain tumors. This brain MRI foundation model also exhibits significant potential for broader applications, including the more brain diseases diagnoses, as well as in the field of MRI generation. The entire pipeline is standardized and modularized, equipped with the capability to interact with experts and engage in continuous learning, thereby enabling iterative optimization of the whole system.

### Brain MRI dataset curation

The development of robust foundation models in neuroimaging critically relies on the availability of large-scale, high-diversity brain MRI datasets. In this study, we have established a comprehensive multi-disease MRI dataset encompassing 24 distinct clinical conditions, including healthy controls, 22 major brain disorders, and miscellaneous neurological diseases (MND) (refer to Glossary 1 for detailed definitions). The brain disease datasets were systematically categorized into five major groups based on their pathological characteristics: (1) Brain Tumor (BT), comprising glioma (GLI), meningioma (MEN), metastasis (MET), and vestibular schwannoma (VS); (2) Cerebrovascular Disease (CVS), including cerebral infarction (CI), intracranial aneurysm (ICA), and intracranial hemorrhage (ICH); (3) Neurodegenerative Disorders (NDD), encompassing Alzheimer’s disease (AD), Parkinson’s disease (PD), multiple sclerosis (MS), frontotemporal dementia (FTD), and amyotrophic lateral sclerosis (ALS); (4) Neuropsychiatric Disorders (NPD), consisting of attention deficit hyperactivity disorder (ADHD), autism spectrum disorder (ASD), major depressive disorder (MDD), schizophrenia (SCZ), post-traumatic stress disorder (PTSD), and cocaine use disorder (CUD); and (5) Other Neurological Conditions, including epilepsy, stroke, traumatic brain injury (TBI), white matter disease (WMD), and MNC (Glossary 1, Fig. 2A). HC and GLI represent the two predominant clinical conditions in our dataset, with HC subjects accounting for 30% of the total cohort and 21.6% of MRI volumes, while GLI cases constitute 18.9% of subjects and 33.5% of MRI volumes (Fig. 2A).

**Fig.2.**
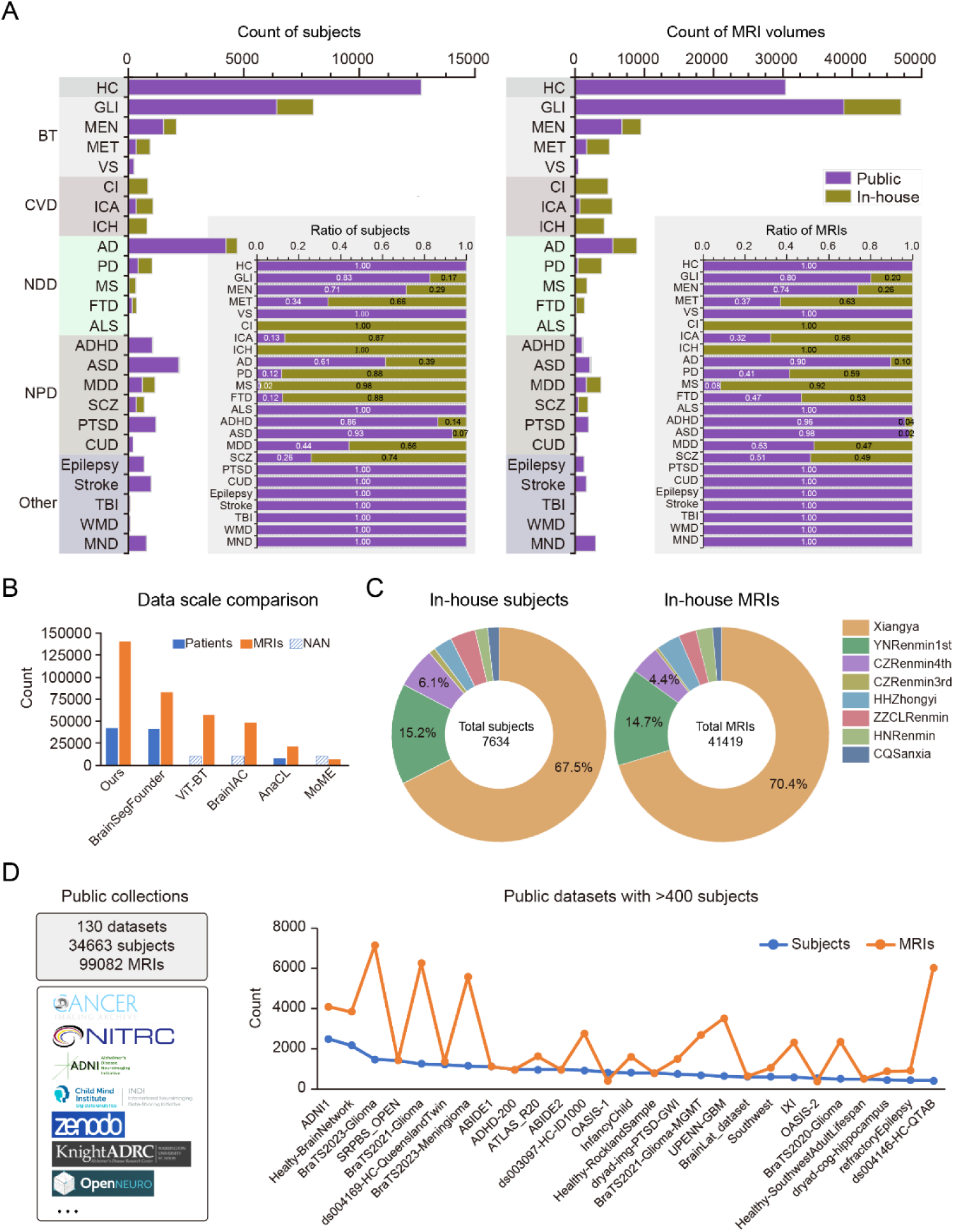
Curation of large-scale brain MRI dataset. (A) Distribution of brain MRI across clinical conditions. Left, distribution of the subjects. Inset panel, the ratio of subjects between public and in-house datasets. Right, distribution of the MRI volumes. Inset panel, the ratio of MRI volumes between public and in-house datasets. (B) Data scale between BrainMRIFM and existing brain MRI foundation models. (C) Distribution of the in-house dataset across clinical centers. Left, distribution of in-house subjects. Right, distribution of inhouse MRI volumes. (D) Curated public brain MRI dataset. Left, summary of the datasets and typical brain MRI sources. Right, distribution of subjects and MRI volumes across various public datasets. Public datasets containing more than 400 subjects are shown.

The curated in-house dataset consisting of 41,419 MRI volumes from 7,634 subjects. Notably, for several critical neurological conditions, including CI, ICA, ICH, PD, MS, FTD, and SCZ, our in-house dataset contributes more than 80% of the total patient cases for each condition. This substantial contribution significantly enhances the disease coverage and diversity of existing brain MRI resources. Furthermore, for MET, MDD, and SCZ, the in-house dataset provides over 50% of the total patient cases, serving as a valuable complement to existing brain MRI collections and substantially expanding the data scale for these conditions (Fig. 2A). To our best knowledge, we curated the largest brain MRI dataset to developing foundation models, holding the potential to sufficiently train MRI foundation models (Fig. 2B). The in-house MRI dataset was systematically collected from eight clinical centers across China, with Xiangya Hospital contributing the majority of cases, representing 67.5% of the total subjects and 70.4% of the MRI volumes (Fig. 2C). The pubic brain MRI dataset was curated from 130 open-source datasets. After pre-processing and manual quality control, the public dataset contains 99,082 MRI volumes from 34,663 subjects. The were 27 datasets contributed significantly to the dataset and each dataset has more than 400 subjects (Fig. 2D). This extensive collection provides a solid and robust base for the development and validation of brain MRI foundation models.

### Slice MRI foundation model

SimMIM-SwinT model was an advanced image foundation model architecture^23^. Based on the pretraining checkpoint of natural images (ImageNet), we re-pretrain it on MRI images and obtained SimMIM-SwinT-based MRI foundation model (Fig. 3A). First, compared to ImageNet foundation model, the MRI foundation model demonstrates a superior ability to extract key features from brain MRI. These features enable better differentiation among the four MRI modalities—T1w, T1ce, T2w, and FLAIR—in t-SNE visualization (Fig. 3B). Second, the MRI foundation model is capable of reconstructing complete images from masked MRI inputs, indicating that the model has effectively learned both global and local MRI features (Fig. 3C). To further validate the high-level semantic representation capabilities of the slice MRI foundation model, we applied it to three conventional brain tumor diagnostic tasks: category recognition (differentiating glioma (GLI), meningioma (MEN), pituitary, and NonTumor) (Fig. 3D), tumor recognition (differentiating NonTumor and GLI) (Fig. 3E), and tumor grading (differentiating LGG and HGG) (Fig. 3F). The MRI foundation model demonstrated superior performance across all tasks compared to models pre-trained on ImageNet. Furthermore, the MRI foundation model achieved robust classification accuracy with only minimal fine-tuning on a small labeled dataset, highlighting its efficiency and generalizability. Furthermore, compared to the foundation models based on the architectures of MoCo v3-ViT^24^ and MAE-ViT^25^. SimMIM-SwinT model exhibited the best performance in all three downstream tasks (Table 1).

**Fig.3.**
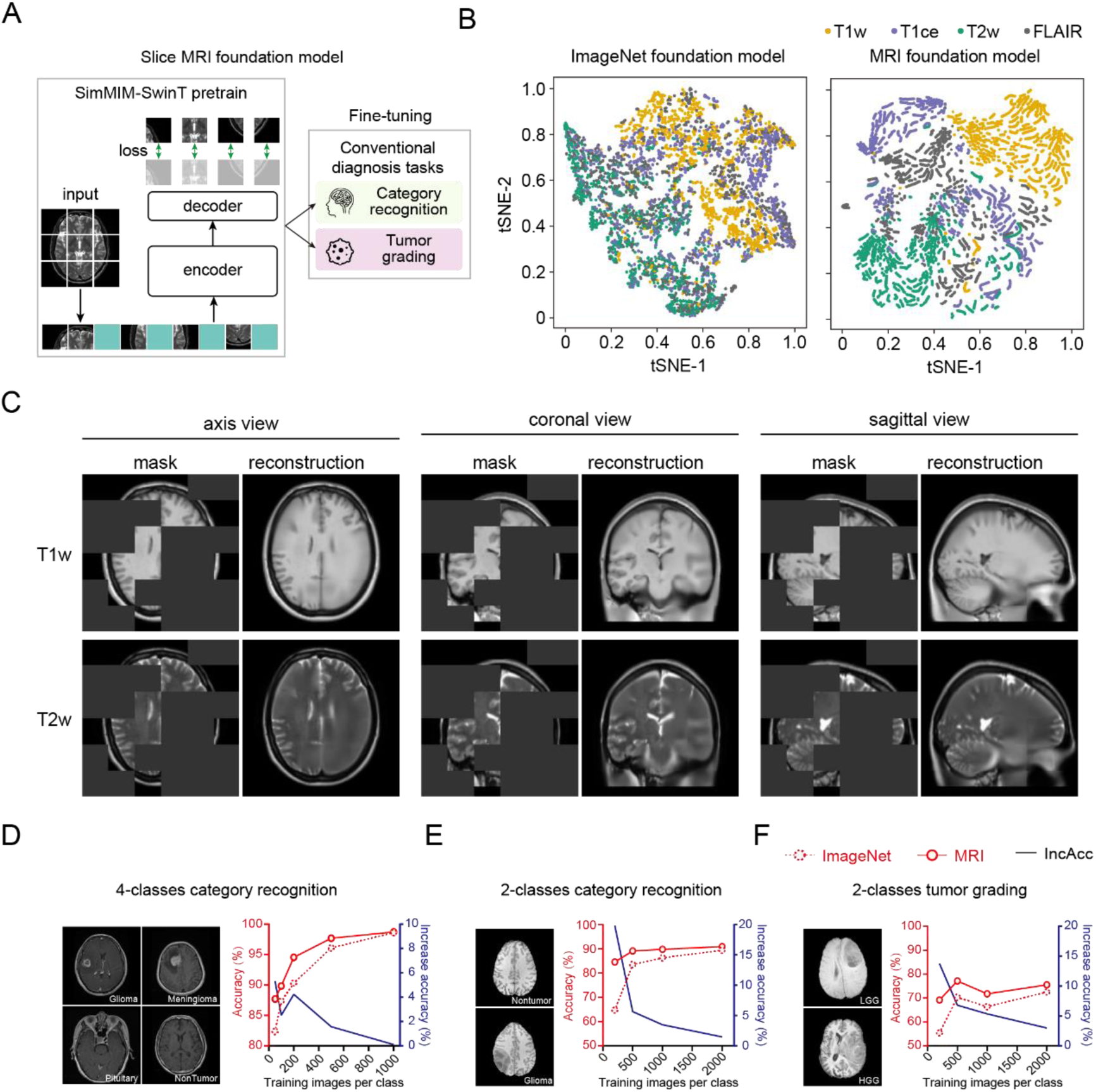
Development and validation of slice MRI foundation models. (A) Pretraining and fine-tuning process of SimMIM-SwinT MRI foundation model. (B) The tSNE visualization of ImageNet and MRI foundation models across four MRI modalities (T1w, T2w, T1ce, and FLAIR). (C) Image reconstruction results of MRI foundation model. The results of two modalities (T1w and T2w) and three views (axis, coronal, and sagittal) are shown. (D) Accuracy of 4-classes (Glioma, Meningioma, Pituitary, and NonTumor) category recognition task. Upper, plot of accuracy of accuracy increases versus training images per class. (E) Accuracy of 2-calsses (NonTumor and Glioma) category recognition task. (F) Accuracy of 2-classes (LGG and HGG) tumor grading task.

**Table 1.** Performance of slice foundation models on three brain tumor diagnostic tasks.

| Foundation models | Accuracy | Accuracy of Brain Tumor Classification (%) | Accuracy of Glioma Classification (%) | Accuracy of Glioma Grading (%) |
| --- | --- | --- | --- | --- |
| MoCo v3-ViT | ImageNet | 93.62 | 74.95 | 65.69 |
|  | MRI | 96.23 | 76.23 | 67.87 |
|  | Increased | 2.61 | 1.28 | 2.18 |
| MAE-ViT | ImageNet | 93.62 | 74.95 | 64.03 |
|  | MRI | 96.00 | 82.91 | 74.52 |
|  | Increased | 2.38 | 7.96 | 10.49 |
| SimMim-SwinT | ImageNet | 98.61 | 89.40 | 72.53 |
|  | MRI | **98.71** | **90.88** | **75.52** |
|  | Increased | 0.10 | 1.47 | 2.99 |

The increased accuracies of MRI foundation models compared to ImageNet foundation models were calculated. MRI foundation models with the best performance for each task are highlighted in bold font.

Herein, the slice MRI foundation model demonstrates the capability to effectively extract critical MRI features, which can not only be directly applied to slice-level diagnostics but also be transferred to volumetric models. This transferability endows the models with the ability to extract and analyze volumetric MRI features, thereby enhancing their diagnostic and clinical utility potential.

### Volume MRI foundation model

We transfer the pretrained parameters of the slice foundation model to the volume foundation model, thereby integrating the respective advantages of slice and volume modeling to establish an MRI foundation model that exhibits high computational efficiency and strong representational capabilities (Fig. 4A). We comprehensively evaluated the effectiveness and generalizability of the volume MRI foundation model across multiple imaging modalities, various brain diseases, and diverse data sources (Fig. 4B). Specifically, we focused on four classical brain imaging modalities (T1w, T2w, T1ce, and FLAIR) to assess the model’s adaptability to different modalities and to explore their diagnostic utility for various diseases. Our investigation encompassed three prevalent brain tumor types, including GLI, MEN, and metastasis (MET), along with their typical subtypes, to validate the model’s capability in handling distinct diagnostic tasks. Furthermore, the adopted six diagnose tasks were derived from highly diverse sources (Table 2), thereby substantiating the model’s generalizability across different data sources.

**Fig.4.**
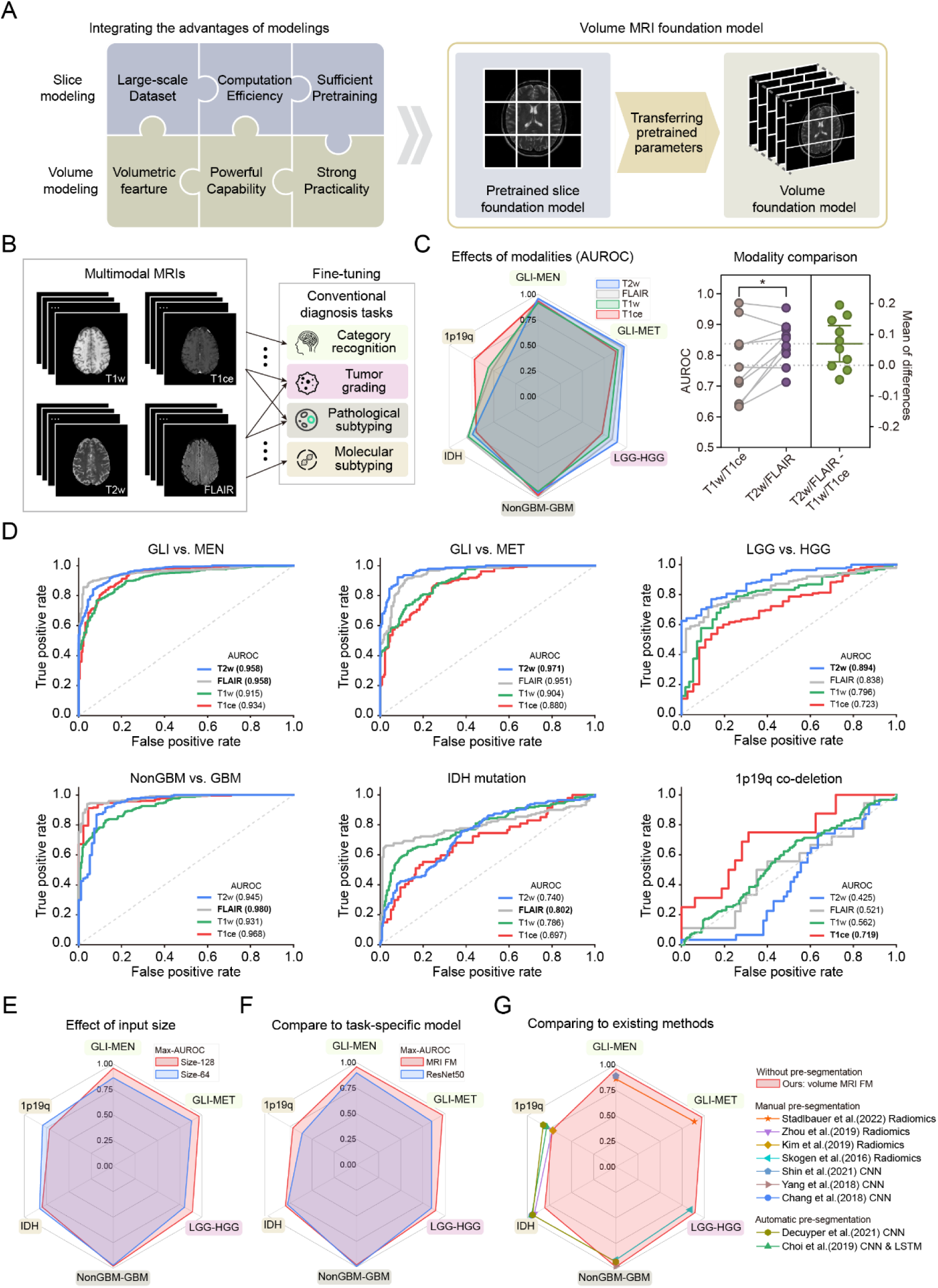
Development and validation of volume MRI foundation models. A) Objectives and processes of developing volume MRI foundation models. Left, objectives that integrating the advantages of slice and volume modeling. Right, transferring the parameters of pretrained slice model to develop volume mode. (B) Experimental settings to validate the performance of volume MRI foundation model. Using multimodal brain MRI scans to achieve multiple conventional diagnosis tasks, including two category recognition tasks (VLI vs. MEN, and GLI vs, MET), one tumor grading task (LGG vs. HGG), one pathological subtyping (NonGBM vs. GBM), and two molecular subtyping tasks (IDH mutation prediction and 1p19q co-deletion prediction). (C) Effect of MRI modalities on SimMIM-SwinT model’s performance (AUROC). Left, radar map of AUROC across six tasks and four MRI modalities. Right, summary graph of AUROC across six tasks between two modality groups (T1w/T1ce, and T2w/FLAIR) (D) Plot of SimMIM-SwinT model’s receiver operating characteristic (ROC) curve across four MRI modalities for each task. (E) Radar map of the maximum AUROCs of SimMIM-SwinT model with different input size. Maximum AUROC for each task is calculated over four MRI modalities. (F) Radar map of AUROCs between SimMIM-SwinT foundation model and task-specific ResNet50 model. (G) Radar map of AUROCs between SimMIM-SwinT foundation model and existing methods for each task.

**Table 2.** Conventional diagnosis tasks in brain tumor and datasets for MRI foundation model evaluation.

| **Task** | **Description** | **Dataset source** |
| --- | --- | --- |
| GLI vs. MEN | Category recognition: distinguishing glioma (GLI) from meningioma (MEN) | ➤ BraTS2023-Glioma<sup>39</sup><br>➤ BraTS2023-Meningioma<sup>40</sup> |
| GLI vs. MET | Category recognition: distinguishing glioma (GLI) from metastasis (MET) | ➤ BraTS2023-Glioma<br>➤ BraTS2023-Metastasis<sup>41</sup> |
| LGG vs. HGG | Tumor grading: distinguishing low-grade (I-II) glioma (LGG) from high-grade (III-IV) glioma (HGG) | ➤ BraTS2020-Glioma<sup>37,38</sup> |
| NonGBM vs. GBM | Pathological subtyping: distinguishing glioblastoma (GBM) from NonGBM subtypes | ➤ TCGA-GBM<sup>42</sup><br>➤ TCGA-LGG<sup>43</sup><br>➤ UPENN-GBM<sup>44,45</sup> |
| IDH | Molecular subtyping: distinguishing IDH-wildtype from IDH-mutant | ➤ TCGA-LGG<br>➤ UPENN-GBM |
| 1p19q | Molecular subtyping: distinguishing 1p19q-wildtype from 1p19q co-deletion | ➤ LGG-1p19qDeletion<sup>46–48</sup><br>➤ TCGA-LGG |

To validate the reliability and interpretability of the model, we investigated the consistency between model characteristics and the clinical prior knowledge. The model demonstrated significantly superior performance on T2w and FLAIR MRIs (AUROC: 0.904 ± 0.079) compared to T1w and T1ce MRIs (AUROC: 0.853 ± 0.090; p < 0.05, paired t-test) across five diagnostic tasks, including GLI vs. MEN, GLI vs. MET, LGG vs. HGG, NonGBM vs. GBM, and IDH-wildtype vs. IDH-mutation (Fig. 4C). From the perspective of imaging characteristics, T2w and FLAIR MRIs primarily capture functional changes in the brain, such as edema and fluid content, while T1w and T1ce MRIs predominantly highlight structural features, including anatomical details and contrast-enhanced regions. This aligns with the clinical practice where T2w and FLAIR MRIs are considered critical for brain tumor diagnosis, as they provide essential information on pathological changes.

The MRI foundation model demonstrated robust performance in tumor category recognition, achieving an overall AUROC of 0.940 ± 0.029 across two sub-tasks and four imaging modalities (Fig. 4D). Specifically, the model attained optimal performance in distinguishing GLI from MEN (AUROC: 0.958) and GLI from MET (AUROC: 0.971) using T2w MRI. In glioma grading tasks, the model achieved an average AUROC of 0.829 ± 0.064 across all modalities, with the highest performance (AUROC: 0.894) observed on T2w MRI (Fig. 4D). For pathological subtyping, the model exhibited strong diagnostic capability, reaching an overall AUROC of 0.961 ± 0.020 across modalities, with FLAIR MRI yielding the best performance (AUROC: 0.980) (Fig. 4D). In molecular subtyping tasks, the model demonstrated significantly superior performance in IDH-mutation recognition (AUROC: 0.765 ± 0.041) compared to 1p19q co-deletion recognition (AUROC: 0.589 ± 0.115) (Fig. 4D). This discrepancy may be attributed to more pronounced radiological feature distinctions between IDH-wildtype and IDH-mutant gliomas, which likely facilitated more accurate classification. To validate the efficiency of our MRI foundation model, we conducted multiple comparisons with conventional deep learning models. Initially, we trained task-specific volume models based on ResNet50 for each diagnostic task (Fig. 4F)^26^. The MRI foundation model demonstrated superior performance across five tasks compared to the ResNet50 models, with nearly identical performance in distinguishing NonGBM from GBM. Compared to most existing models, the MRI foundation model offers advantages including task generality, streamlined processes, and superior performance across most tasks (Fig. 4G) ^27–35^. It operates directly on raw MRI sequences without pre-segmentation, whereas other approaches typically require manual or automatic tumor segmentation. The MRI foundation model outperforms the majority of existing methods in tasks such as category recognition, tumor grading, and pathological subtyping. However, it shows slightly inferior performance in molecular subtyping tasks compared to some existing methods. This discrepancy may be attributed to the fact that molecular subtyping relies on fine-grained features within the tumor region, and pre-segmentation of tumors may enhance the identification of such detailed characteristics.

To elucidate the interpretability of the MRI foundation model, we employed Gradient-weighted class activation mapping (Grad-CAM) to visualize the critical regions of interest identified by the model. The generated heatmaps demonstrated that the model consistently localized tumor-related regions with high precision across all five diagnostic tasks (Fig. S1). Notably, the volumetric MRI foundation model exhibited robust spatial awareness, not only accurately identifying tumor-relevant slices along the MRI sequence dimension but also precisely focusing on tumor-specific areas within individual slices. The foundation model demonstrates superior feature representation capabilities, facilitating accurate lesion localization and comprehensive characterization across a spectrum of tumor sizes, spatial distributions, and imaging signatures in various brain tumor subtypes. The foundational model’s characterization of MRI modalities and tumor region features aligns closely with established clinical expertise, thereby enhancing its interpretability and underscoring its practical relevance in clinical applications.

## Discussion

This work presents two pivotal advances for brain MRI foundation models: (1) BrainMRIFM, a computationally efficient slice-to-volume pipeline that overcomes dimensional constraints in MRI dataset and modeling processes, and (2) the largest multi-modal neuroimaging dataset to date (140,501 volumes from 42,297 subjects), addressing critical gaps in scale and diversity for robust pretraining.

A critical challenge in developing foundation models for brain MRI lies in aligning data scales with model architectures. Each MRI volume can yield dozens to hundreds of MRI slices, resulting in a substantially larger slice-level dataset that facilitates robust pretraining of slice foundation models. This scale advantage is compounded by the significantly reduced parameter size and computational demands of slice models, which enable more comprehensive pretraining for efficient MRI feature extraction. Representative approaches, such as BME-X, have demonstrated the feasibility of slice MRI foundation model^20^。 However, since brain MRIs are intrinsically volumetric, 3D and 2.5D volume models hold greater clinical potential. 3D models (e.g., BrainIAC, AnaCL, BrainSegFounder, MoME)^10–15^ employ 3D convolutions or 3D tokenizers to directly capture spatial dependencies ; however, they face significant computational complexity and insufficient pretraining due to the scarcity of medical data. In contrast, 2.5D (video-style) models mimic radiologists’ slice-by-slice analysis by first extracting slice-level features and then aggregating them volumetrically. Our BrainMRIFM framework introduces a computationally efficient slice-to-volume (2.5D) pipeline that synergizes the advantages of slice modeling and volume modeling. BrainMRIFM simultaneously achieves both computational efficiency and powerful representational capability, demonstrating great potential for practical clinical applications.

We have curated an unprecedented brain MRI dataset that serves as a robust foundation for developing brain MRI foundation models. In comparison to BrainSegFounder, which previously utilized the largest brain MRI dataset, BrainMRIFM contains 1.7 times more MRI scans (∼140K in BrainMRIFM versus ∼83K in BrainSegFounder)^12^. Importantly, while BrainSegFounder’s data originated from limited sources (UK Biobank, BraTS challenge, and ATLAS v2.0 datasets), BrainMRIFM aggregates data from 130 public datasets and 8 clinical centers across 24 distinct clinical conditions. This demonstrates a significantly greater diversity, enabling the foundation model to learn more comprehensive features and achieve improved generalization capabilities. The BrainIAC MRI foundation model was also developed using 34 brain MRI datasets, but it contains significantly fewer MRI scans (∼50K) compared to BrainMRIFM (∼140K)^10^. Furthermore, we have established innovative and distinctive in-house brain MRI datasets that complement existing open-source data by including both common and rare but clinically significant brain diseases. Publicly available MRIs for CI, ICA, ICH, PD, MS, FTD, and SCZ remain scarce, severely limiting deep learning researches in these areas. We have strategically collected MRI data, with our in-house dataset contributing over 80% of the total patient cases for these diseases. This substantial addition dramatically enhances the disease coverage and diversity of current brain MRI resources. The unprecedented brain MRI dataset not only increases the scale and diversity of the data but also significantly expands the potential applications of BrainMRIFM.

The BrainMRIFM framework presents several opportunities for further optimization. Currently, the pretraining strategy does not fully utilize the unique characteristics of brain MRI. Incorporating domain-specific clinical priors during pretraining could potentially enhance the model’s performance. Additionally, the inherent imbalance in the MRI dataset may restrict the model’s ability to effectively capture pathological features from conditions with limited MRI scans. Strategic approaches, such as weighting for rare disease categories or targeted re-pretraining using disease-specific datasets, could improve both the generalizability and robustness of the foundation models.

## Methods

### Brain MRI dataset curation and preprocessing

We have exhausted almost all the available open-source brain MRI imaging, including datasets from TCIA, NITRC, ADNI, INDI, ZENODO, OpenNeuro. The custom brain MRI datasets were collected from 8 hospitals in China: Xiangya Hospital of Central South University (Xiangya), The First People’s Hospital of Yunnan Province (YNRenmin1st), The Third People’s Hospital of Chenzhou (CZRenmin3rd), The Fourth People’s Hospital of Chenzhou (CZRenmin4th), Huaihua Traditional Chinese Medicine Hospital (HHZhongyi), Chaling County People’s Hospital of Zhuzhou (ZZCLRenmin), Hunan Provincial People’s Hospital (HNRenmin), and Chongqing University Three Gorges Hospital (CQSanxia).

All brain MRI dataset are processed followed the standard pipeline as follows: 1) Establish the MRI preprocessing pipeline using Nipype. 2) Convert all the source data into NIfTI file using Nibabel and save the data in the uint32 format. 3) Implement N4 bias field correction using ANTs. 4) Adjust the voxel resolution to 1×1×1 mm^3^ using ANTs. 5) Implement 3D MRI registration using the FSL tool and the MNI-152 brain atlas. 6) Remap the values of each 3D MRI data to 0-255 and save multiple images for manual quality inspection using Nibabel. 7) Extract axial, coronal, and sagittal slices from 3D MRI. Considering the diversity of image quality, we adopt the perception-based image quality estimator (PIQE) to control the quality of MRI slices (Fig. S2). The dataset for pretraining slice foundation model only included MRI slices that met the PIQE threshold (PIQE ≤ 60).

### Downstream diagnostic tasks

We evaluated slice MRI foundation models on three downstream diagnostic tasks, including Brain Tumor Classification, Glioma Classification, and Glioma Grading. Brain Tumor Classification uses a dataset that contains 7,023 brain MRI images and classifies each image into 4 classes: glioma, meningioma, pituitary, and nontumor^36^. The ratio of MRI images in the training and testing sets is 4:1 for each class. Glioma Classification task is based on Brain Tumor Segmentation Challenge 2020 (BraTS2020), which consists of brain MRIs and brain tumor segmentation results from patients with glioblastoma or high grade glioma (GBM/HGG) and lower grade glioma (LGG)^37,38^. For each MRI slice, “glioma” refers to slices with tumor, while “nontumor” refers to slices without the tumor. Based on the settings of the glioma classification task, the glioma grading task further classifies glioma slices into LGG or HGG. The grade of MRI scan was identified in BraTS2020, and the grade of glioma slices was derived from the volume MRI. We evaluated volume MRI foundation models on six downstream diagnostic tasks which were derived from highly diverse sources (Table 2), thereby substantiating the model’s generalizability across different data sources.

All experiments were conducted on the Biomedical Computing Platform of National Biomedical Imaging Center, Peking University. We trained the foundational models using OpenMMLab tool on a high-performance computing cluster equipped with 8× NVIDIA A800 GPUs. For each downstream task, the training and test datasets were randomly divided 4:1 at the patient level.

## Data Availability

All data produced in the present study are available upon reasonable request to the authors

## Acknowledgements

We thank Rui Ma (Peking University) for technical support on the high-performance computing cluster. We thank the Biomedical Computing Platform of National Biomedical Imaging Center at Peking University for computational resources. This work is supported by Beijing Natural Science Foundation (7254542) and China Postdoctoral Science Foundation (8206301107).

## Figures and Tables

**Glossary 1.** Terminology of brain clinical conditions.

| Term | Expansion | Clinical category | Description |
| --- | --- | --- | --- |
| HC | Healthy control | HC | Subjects with normal cognition and without determined brain disease. |
| GLI | Glioma | BT | Brain tumor originating from glial cells. |
| MEN | Meningioma | BT | Benign tumor arising from the meninges. |
| MET | Metastasis | BT | Tumor originated from other site and spread to brain. |
| VS | Vestibular schwannoma | BT | Benign tumor affecting the nerve responsible for balance and hearing. |
| CI | Cerebral infarction | CVD | Brain tissue damage caused by a lack of blood flow. |
| ICA | Intracranial aneurysm | CVD | A weakened blood vessel in the brain that bulges and may rupture. |
| ICH | Intracranial hemorrhage | CVD | Bleeding within the skull, which can occur in the brain or surrounding areas. |
| AD | Alzheimer's disease | NDD | Characterized by memory loss and cognitive decline. |
| PD | Parkinson's disease | NDD | Characterized by tremors, rigidity, and movement difficulties. |
| MS | Multiple sclerosis | NDD | An autoimmune disease damaging the protective covering of nerves, disrupting communication between the brain and body. |
| FTD | Frontotemporal dementia | NDD | Causing progressive damage to the frontal and temporal lobes, leading to behavioral and language changes. |
| ALS | Amyotrophic lateral sclerosis | NDD | Affecting motor neurons, leading to muscle weakness and paralysis. |
| ADHD | Attention-deficit/hyperactivity disorder | NPD | Characterized by inattention, hyperactivity, and impulsivity. |
| ASD | Autism spectrum disorder | NPD | Affecting communication, behavior, and social interaction. |
| MDD | Major depressive disorder | NPD | Characterized by persistent sadness and loss of interest. |
| SCZ | Schizophrenia | NPD | Characterized by distorted thinking, perceptions, and emotional responses. |
| PTSD | Post-Traumatic stress disorder | NPD | A mental health condition triggered by traumatic events, causing flashbacks and severe anxiety. |
| CUD | Cocaine use disorder | NPD | Characterized by involving addiction to cocaine. |
| Epilepsy | Epilepsy | Other | Characterized by recurrent seizures. |
| Stroke | Stroke | Other | A sudden interruption of blood flow to the brain, causing cell death. |
| TBI | Traumatic brain injury | Other | Brain damage caused by an external force, often resulting in cognitive or physical impairment. |
| WMD | White matter damage | Other | Injury to the brain's white matter, affecting communication between brain regions. |
| MND | Miscellaneous neurological diseases | Other | Brain-related neurological conditions not specifically listed. |

BT: brain tumor. CVD: cerebrovascular disease. NDD: neurodegenerative disorder. NPD: neuropsychiatric disorder.

**Fig. S1.**
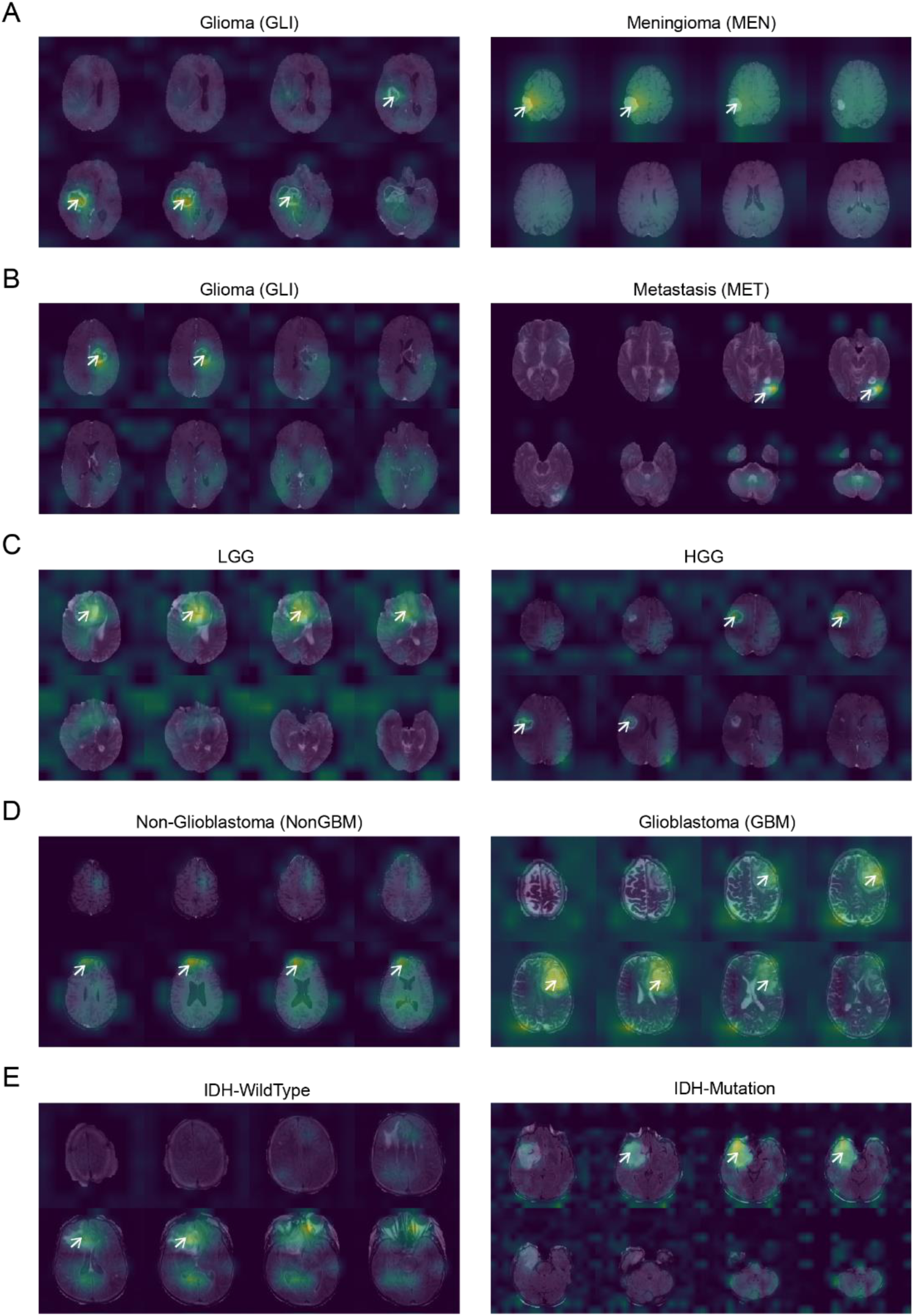
Saliency visualization of SimMIM-SwinT volume foundation model over five tasks. White arrows mark lesions accurately located by the model. (A) Attention maps for GLI vs. MEN task. (B) Attention maps for GLI vs. MET task. (C) Attention maps for LGG vs. HGG task. (D) Attention maps for NonGBM vs. GBM task. (E) Attention maps for IDH mutation prediction task.

**Fig. S2.**
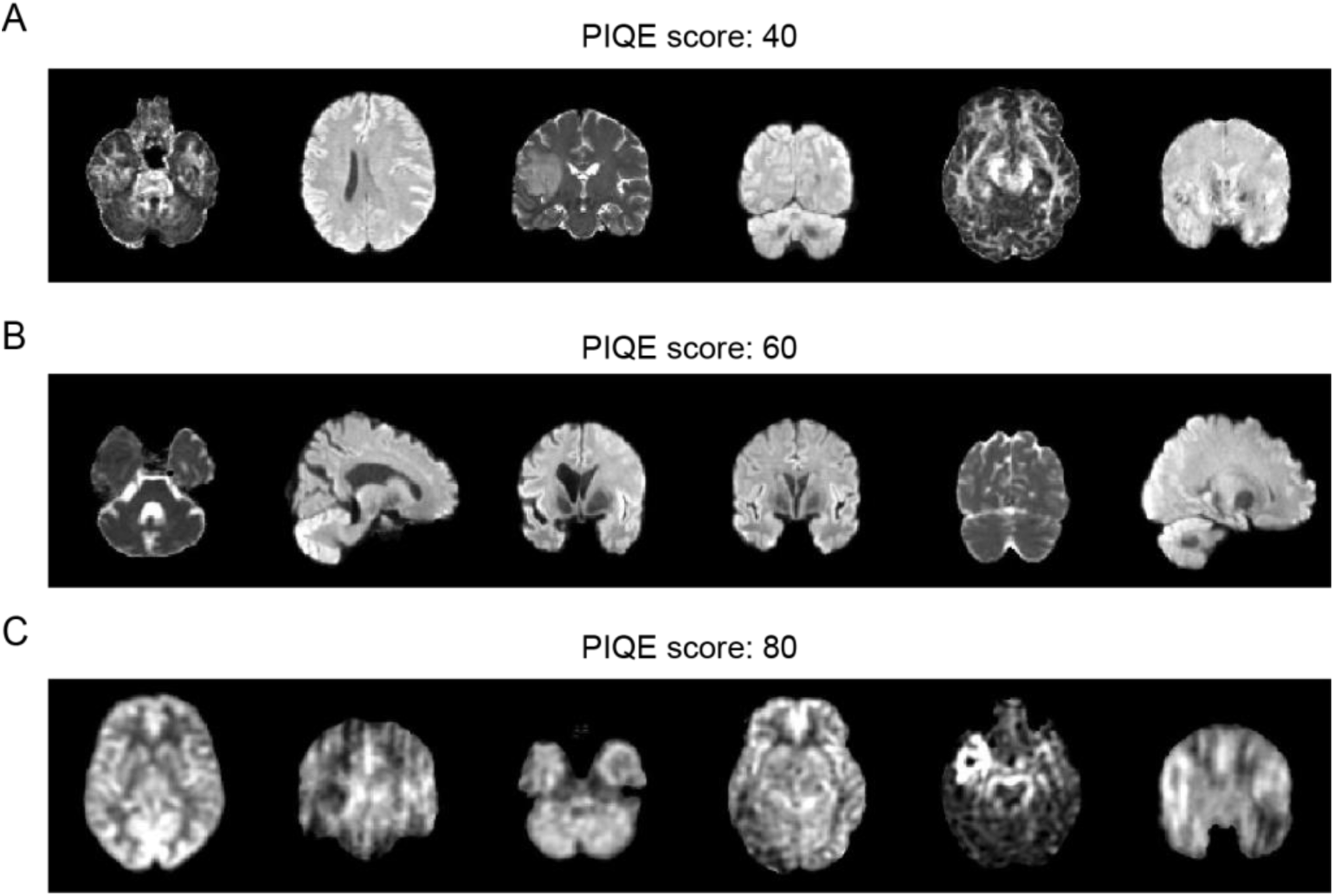
MRI slices with different PIQE score. (A) PIQE = 40. (B) PIQE = 60. (C) PIQE = 80. For each PIQE setting, six MRI slices are shown.

## References

1. Sebenius, I. et al. Robust estimation of cortical similarity networks from brain MRI. Nat. Neurosci. 1–11 (2023) doi:10.1038/s41593-023-01376-7.

2. Bethlehem, R. A. I. et al. Brain charts for the human lifespan. Nature 604, 525–533 (2022).

3. Feinberg, D. A. et al. Next-generation MRI scanner designed for ultra-high-resolution human brain imaging at 7 Tesla. Nat. Methods 20, 2048–2057 (2023).

4. Rudie, J. D., Rauschecker, A. M., Bryan, R. N., Davatzikos, C. & Mohan, S. Emerging Applications of Artificial Intelligence in Neuro-Oncology. Radiology 290, 607–618 (2019).

5. Lee, J. et al. Deep learning-based brain age prediction in normal aging and dementia. Nat. Aging 2, 412–424 (2022).

6. Moor, M. et al. Foundation models for generalist medical artificial intelligence. Nature 616, 259–265 (2023).

7. Pai, S. et al. Foundation model for cancer imaging biomarkers. Nat. Mach. Intell. 6, 354–367 (2024).

8. Ying, H. et al. A multicenter clinical AI system study for detection and diagnosis of focal liver lesions. Nat. Commun. 15, 1131 (2024).

9. Wang, Y.-R. et al. Screening and diagnosis of cardiovascular disease using artificial intelligence-enabled cardiac magnetic resonance imaging. Nat. Med. 30, 1471– 1480 (2024).

10. Tak, D. et al. A foundation model for generalized brain MRI analysis. 2024.12.02.24317992 Preprint at 10.1101/2024.12.02.24317992 (2024).

11. Barbano, C. A., Brunello, M., Dufumier, B. & Grangetto, M. Anatomical Foundation Models for Brain MRIs. Preprint at http://arxiv.org/abs/2408.07079 (2024).

12. Hatamizadeh, A., et al. Swin UNETR: Swin Transformers for Semantic Segmentation of Brain Tumors in MRI Images. Preprint at 10.48550/arXiv.2201.01266 (2022).

13. Cox, J. et al. BrainSegFounder: Towards 3D foundation models for neuroimage segmentation. Med. Image Anal. 97, 103301 (2024).

14. Zhang, X. et al. A Foundation Model for Brain Lesion Segmentation with Mixture of Modality Experts. in Medical Image Computing and Computer Assisted Intervention – MICCAI 2024 (eds. Linguraru, M. G. et al.) 379–389 (Cham, 2024). doi:10.1007/978-3-031-72390-2_36.

15. Chen, M. et al. Medical image foundation models in assisting diagnosis of brain tumors: a pilot study. Eur. Radiol. (2024) doi:10.1007/s00330-024-10728-1.

16. Liu, H. et al. M3AE: Multimodal Representation Learning for Brain Tumor Segmentation with Missing Modalities. Preprint at http://arxiv.org/abs/2303.05302 (2023).

17. Wang, W. et al. FreMIM: Fourier Transform Meets Masked Image Modeling for Medical Image Segmentation. Preprint at http://arxiv.org/abs/2304.10864 (2023).

18. Wang, K. et al. Improving brain tumor segmentation with anatomical prior-informed pre-training. Front. Med. 10, 1211800 (2023).

19. Prabhakar, C. et al. ViT-AE++: Improving Vision Transformer Autoencoder for Self-supervised Medical Image Representations. Preprint at http://arxiv.org/abs/2301.07382 (2023).

20. Sun, Y., Wang, L., Li, G., Lin, W. & Wang, L. A foundation model for enhancing magnetic resonance images and downstream segmentation, registration and diagnostic tasks. Nat. Biomed. Eng. 1–18 (2024) doi:10.1038/s41551-024-01283-7.

21. Ma, J. et al. Segment anything in medical images. Nat. Commun. 15, 654 (2024).

22. Putz, F. et al. The Segment Anything foundation model achieves favorable brain tumor auto-segmentation accuracy in MRI to support radiotherapy treatment planning. Strahlenther. Onkol. 201, 255–265 (2025).

23. Xie, Z. et al. SimMIM: a Simple Framework for Masked Image Modeling. in 2022 IEEE/CVF Conference on Computer Vision and Pattern Recognition (CVPR) 9643–9653 (IEEE, New Orleans, LA, USA, 2022). doi:10.1109/CVPR52688.2022.00943.

24. He, K., Fan, H., Wu, Y., Xie, S. & Girshick, R. Momentum Contrast for Unsupervised Visual Representation Learning. Preprint at 10.48550/arXiv.1911.05722 (2020).

25. He, K., et al. Masked Autoencoders Are Scalable Vision Learners. Preprint at 10.48550/arXiv.2111.06377 (2021).

26. Wang, X., Girshick, R., Gupta, A. & He, K. Non-local Neural Networks. in 2018 IEEE/CVF Conference on Computer Vision and Pattern Recognition 7794–7803 (2018). doi:10.1109/CVPR.2018.00813.

27. Stadlbauer, A. et al. Radiophysiomics: Brain Tumors Classification by Machine Learning and Physiological MRI Data. Cancers 14, 2363 (2022).

28. Zhou, H. et al. Machine learning reveals multimodal MRI patterns predictive of isocitrate dehydrogenase and 1p/19q status in diffuse low- and high-grade gliomas. J. Neurooncol. 142, 299–307 (2019).

29. Kim, D. et al. Prediction of 1p/19q Codeletion in Diffuse Glioma Patients Using Pre-operative Multiparametric Magnetic Resonance Imaging. Front. Comput. Neurosci. 13, 52 (2019).

30. Skogen, K. et al. Diagnostic performance of texture analysis on MRI in grading cerebral gliomas. Eur. J. Radiol. 85, 824–829 (2016).

31. Shin, I. et al. Development and Validation of a Deep Learning–Based Model to Distinguish Glioblastoma from Solitary Brain Metastasis Using Conventional MR Images. Am. J. Neuroradiol. 42, 838–844 (2021).

32. Yang, Y. et al. Glioma Grading on Conventional MR Images: A Deep Learning Study With Transfer Learning. Front. Neurosci. 12, 804 (2018).

33. Chang, K. et al. Residual Convolutional Neural Network for the Determination of *IDH* Status in Low- and High-Grade Gliomas from MR Imaging. Clin. Cancer Res. 24, 1073–1081 (2018).

34. Decuyper, M., Bonte, S., Deblaere, K. & Van Holen, R. Automated MRI based pipeline for segmentation and prediction of grade, IDH mutation and 1p19q co-deletion in glioma. Comput. Med. Imaging Graph. Off. J. Comput. Med. Imaging Soc. 88, 101831 (2021).

35. Choi, K. S., Choi, S. H. & Jeong, B. Prediction of IDH genotype in gliomas with dynamic susceptibility contrast perfusion MR imaging using an explainable recurrent neural network. Neuro-Oncol. 21, 1197–1209 (2019).

36. Masoud, N. Brain Tumor MRI Dataset. Kaggle.

37. Bakas, S. et al. Advancing The Cancer Genome Atlas glioma MRI collections with expert segmentation labels and radiomic features. Sci. Data 4, 170117 (2017).

38. Menze, B. H. et al. The Multimodal Brain Tumor Image Segmentation Benchmark (BRATS). IEEE Trans Med Imaging 34, 1993–2024 (2015).

39. Kazerooni, A. F. et al. The Brain Tumor Segmentation (BraTS) Challenge 2023: Focus on Pediatrics (CBTN-CONNECT-DIPGR-ASNR-MICCAI BraTS-PEDs). Preprint at 10.48550/arXiv.2305.17033 (2024).

40. LaBella, D. et al. The ASNR-MICCAI Brain Tumor Segmentation (BraTS) Challenge 2023: Intracranial Meningioma. Preprint at 10.48550/arXiv.2305.07642 (2023).

41. Moawad, A. W. et al. The Brain Tumor Segmentation (BraTS-METS) Challenge 2023: Brain Metastasis Segmentation on Pre-treatment MRI. Preprint at 10.48550/arXiv.2306.00838 (2024).

42. Scarpace, L. et al. The Cancer Genome Atlas Glioblastoma Multiforme Collection (TCGA-GBM). The Cancer Imaging Archive 10.7937/K9/TCIA.2016.RNYFUYE9 (2016).

43. Pedano, N. et al. The Cancer Genome Atlas Low Grade Glioma Collection (TCGA-LGG). The Cancer Imaging Archive 10.7937/K9/TCIA.2016.L4LTD3TK (2016).

44. Bakas, S. et al. Multi-parametric magnetic resonance imaging (mpMRI) scans for de novo Glioblastoma (GBM) patients from the University of Pennsylvania Health System (UPENN-GBM). The Cancer Imaging Archive 10.7937/TCIA.709X-DN49 (2021).

45. Bakas, S., et al. The University of Pennsylvania glioblastoma (UPenn-GBM) cohort: advanced MRI, clinical, genomics, & radiomics. Sci. Data 9, 453 (2022).

46. Erickson, B., Akkus, Z., Sedlar, J. & Korfiatis, P. Data from LGG-1p19qDeletion. The Cancer Imaging Archive 10.7937/K9/TCIA.2017.DWEHTZ9V (2017).

47. Akkus, Z. et al. Predicting Deletion of Chromosomal Arms 1p/19q in Low-Grade Gliomas from MR Images Using Machine Intelligence. J. Digit. Imaging 30, 469– 476 (2017).

48. Erickson, B. J., Korfiatis, P., Akkus, Z., Kline, T. & Philbrick, K. Toolkits and Libraries for Deep Learning. J. Digit. Imaging 30, 400–405 (2017).

